# Protocol for an Observational Prospective Study Linking RDoC-Based Phenotypes with Clinical and Care-Related Outcomes: The VeRDoC – Study

**DOI:** 10.1101/2025.06.04.25328893

**Authors:** Kristin Koller-Schlaud, Johannes Meixner, Kerstin Jost, Frauke Waghals, Johannes Rentzsch, Bernd R. Förstner, Martin Heinze, Joachim Behr, Michael A. Rapp, Mira Tschorn

**Affiliations:** Department of Psychiatry, Psychotherapy and Psychosomatics, University Hospital Ruppin-Brandenburg, Brandenburg Medical School, Neuruppin, Germany; Department of Psychology, Brandenburg Medical School, Neuruppin, Germany; Center for Mental Health, Immanuel Clinic Rüdersdorf, Brandenburg Medical School, Rüdersdorf bei Berlin, Germany; Social and Preventive Medicine, Department of Sports and Health Sciences, University of Potsdam, Potsdam, Germany; Department of Psychiatry and Psychotherapy, Charité - Universitätsmedizin Berlin, Berlin, Germany; Faculty of Health Sciences Brandenburg, Brandenburg Medical School Theodor Fontane, Germany; German Center for Mental Health (DZPG), partner site Berlin/Potsdam, Germany

**Keywords:** research domain criteria (RDoC), transdiagnostic psychiatry, care-related outcomes, multimethod approach, ageing

## Abstract

**Introduction:** The Research Domain Criteria (RDoC) approach initiated by the National Institute of Mental Health provides a comprehensive framework for guiding research on mental illness and health. Since retrospective studies have indicated associations between RDoC characteristics and clinically relevant as well as care-relevant outcomes, there is a need for prospective, theory-driven investigations that systematically link *a priori* defined assessments of RDoC constructs to clinically and care-relevant outcomes in a transdiagnostic psychiatric sample.

**Methods and Analysis:** This prospective observational study assesses six domains - Positive Valence Systems, Negative Valence Systems, Cognitive Systems, Social Processes, Arousal and Regulatory Systems, and Sensorimotor Systems employing a comprehensive set of self-report and additional paradigms to assess cognitive functioning developed a priori in alignment with the Research Domain Criteria framework while also assessing clinically and care-relevant variables (e.g., length of hospital stay). A total of 300 adult participants will be recruited among in- and outpatients of two psychiatric hospitals in Germany (patient group). Including healthy individuals will allow for the investigation of continuous variations in psychological functioning rather than categorical distinctions between health and disease. Data collection includes self-reports, clinician ratings, file review, and behavioral assessments. Electroencephalography (EEG) is recorded in a subgroup of participants. A confirmatory factor analysis will be conducted to reproduce the factor structure and regression models will be used to investigate associations between RDoC domains and clinically relevant as well as care-related variables.

**Ethics and dissemination:** Ethics approval was obtained from the local ethics committee of the Brandenburg Medical School – Theodor Fontane (E-01-20220822). Results will be disseminated through peer-reviewed journals and academic conferences.

**Article Summary:** 

**Strengths and limitations of this study:**

- The study employs a set of assessments explicitly designed a priori in alignment with the RDoC framework.
- The study employs a dimensional approach to psychological functioning that cuts across traditional psychiatric diagnoses and investigates a transdiagnostic psychiatric patient sample.
- The study addresses a key challenge of enhancing translational value by integrating indicators of service use, functional outcomes, and quality of life, as well as by incorporating external validators.
- The cross-sectional design limits the ability to draw causal inferences about the relationship between RDoC domains and clinically relevant and care-related variables.
- The limited multi-method approach with reliance on mostly self-report and cognitive paradigms may be influenced by subjective bias or situational factors.

## 1. Introduction

The Research Domain Criteria (RDoC) initiative, launched by the National Institute of Mental Health (NIMH) in 2009, provides a comprehensive framework for conceptually organizing and guiding psychobiological research on mental illness and health. It was developed in response to growing evidence regarding reduced clinical validity and utility of categorical diagnostic approaches, which have been accompanied by inconsistent findings across contemporary research methodologies. The goal was to promote a new, broadly conceived dimensional concept of mental illness, grounded in varying degrees of dysfunction across psychological and neurobiological systems, with the hope that research within the RDoC framework would increase the validity and clinical utility of characterizations, inform revisions of current diagnostic categories, and ultimately improve prevention and intervention efforts [1].

The RDoC approach adopts a lifespan perspective, aiming to understand mental states across developmental stages while integrating biological, physiological, and behavioral measures, and by linking neurobiological findings with psychopathology to enhance diagnosis, treatment, and prevention strategies [2]. The framework is organized as a matrix that groups constructs and subconstructs of basic human neurobehavioral functioning into six domains: Positive Valence Systems (PVS), Negative Valence Systems (NVS), Cognitive Systems (CoS), Social Processes (SoP), Arousal and Regulatory Systems (ARS), and Sensorimotor Systems (SmS) [1]. Each domain is arranged hierarchically and mapped onto seven units of analysis: molecules, cells, circuits, physiology, behavior, self-report, and paradigms. Briefly summarized, the PVS domain includes mechanisms associated with processing and responding to positive motivational situations, such as reward responsiveness and reward learning. The NVS domain encompasses responses to aversive situations, including acute and potential threats, loss, and aggression. The CoS domain addresses cognitive functions such as attention, perception, memory, language, and cognitive control. The SoP domain involves processes of affiliation, attachment, social communication, and the perception and understanding of self and others. The ARS domain can be subdivided into three constructs and includes arousal, circadian rhythms, and sleep/wakefulness. The SmS domain addresses control and execution of motor behaviors and includes constructs such as motor action, agency and ownership, and habit.

Importantly, RDoC highlights the influence of both lifespan development and environmental factors as crucial context variables for understanding neurobehavioral mechanisms [3, 4]. Research suggests that the expression of psychological mechanisms can vary across different stages of life; for example, cognitive control within the CoS domain continues to mature into early adulthood and declines in older age [5]. Age-related differences in threat processing further illustrate this principle: Older adults exhibit heightened attention bias toward physical threats, moderated by cognitive control capacity, which has implications for anxiety mechanisms across the lifespan [6].

As the ultimate goal of the RDoC initiative is to positively impact clinical outcomes through the identification of transdiagnostic, brain-behavior constructs and the development of dimensional, integrative analytic approaches [4]; [7], it is crucial to investigate how RDoC domains relate to clinical and care-related parameters. These include not only psychiatric symptoms and diagnostic information, but also service use, functional outcomes, and quality of life measures.

Previous retrospective studies have already indicated associations between RDoC characteristics and care-relevant outcomes. For instance, a retrospective analysis of inpatient psychiatric data found that dysfunctions within the cognitive systems and arousal/regulatory systems domains were associated with longer treatment durations [8]. Further work from the same group demonstrated that features within the negative valence and social processes domains predicted the risk of readmission following inpatient discharge [8]. Follow-up studies showed that improvements in negative valence system scores during hospitalization were associated with reduced readmission risk [9]. Additionally, RDoC constructs have been shown to more accurately predict transdiagnostic phenomena, such as suicidality, than traditional categorical diagnoses, such as the presence of a depressive disorder [10]).

Furthermore, there is growing evidence from studies investigating the relationship between RDoC constructs and quality-of-life outcomes. For example, Verdejo-García et al. [11] identified internalizing transdiagnostic profiles characterized by high behavioral activation and low inhibitory control, which were associated with lower quality of life. Similarly, Diamond [12] summarized evidence showing that better executive functioning, a key construct within the RDoC Cognitive Systems domain, predicts higher quality of life across the lifespan. Additionally, Whitton et al. [13] demonstrated that anhedonia and its neural correlates within the Positive Valence Systems domain are robustly linked to quality-of-life outcomes in individuals with mood disorders, both cross-sectionally and longitudinally.

The present study aims to systematically assess core domains of psychological functioning in a transdiagnostic psychiatric clinical sample using a set of self-report, cognitive paradigms, and physiological (EEG-based) assessments explicitly developed in alignment with the RDoC framework. Furthermore, the study seeks to investigate the associations between RDoC domains and clinically relevant and care-related variables, and to examine potential effects of age on these associations in our transdiagnostic psychiatric sample.

To this end, we utilize self-reported indicators of core psychological processes that are hypothesized to reflect underlying biological mechanisms, aiming to build integrative biopsychological models of clinical symptomatology. Extending previous work that conducted post-hoc mappings of traditional assessments onto RDoC domains (e.g., [14]), we designed our assessment battery *a priori*, guided systematically by RDoC constructs. This methodological strength enables a more systematic and theoretically grounded investigation of RDoC domains. Furthermore, we include clinical outcome ratings as additional external validators. This approach follows recommendations in the literature emphasizing the importance of linking RDoC constructs to clinically relevant variables to enhance translational value [15].

Given that we designed our study *a priori* on the basis of the RDoC framework and in line with its transdiagnostic approach, participant inclusion in our study will not be limited to specific diagnostic categories, as was the case in some earlier post-hoc mapping studies. Instead, we aim to capture a broad spectrum of psychopathology by recruiting participants from a general psychiatric sample as well as including a control sample.

Specifically, we aim to test the hypothesis that selected RDoC constructs are significantly linked to psychological burden, while accounting for sociodemographic, socioeconomic, and age effects. Moreover, we plan to analyze whether RDoC domains account for additional variance when predicting clinically relevant (e.g., suicidality, symptom severity) and care-related outcomes (e.g., length of hospital stay, readmission) beyond the variance explained by overall illness severity and traditional diagnostic categories. Further, we intend to analyze in how far RDoC domains can predict changes in clinical severity.

The long-term goal of our research is to contribute to the development of precision medicine approaches in mental health care by identifying novel treatment targets based on comprehensive and dimensional approaches of psychopathology, enabling more personalized treatment selection, and ultimately optimizing psychiatric care.

## 2. Methods and Analysis

### 2.1 Study Design

This study is prospective, observational and multicentric with two participating psychiatric hospitals. To minimize participant burden and optimize recruitment procedures, this study project was combined with a separate study project that also focuses on a transdiagnostic approach within the RDoC framework. For the purposes of this study protocol, the first project is referred to as the “main study”, while the second is referred to as the “EEG-substudy”. The main study assesses the six current RDoC domains and examines the clinical relevance of these domains using external care-related and clinical outcomes as validators. The second study addresses cognitive control within the CoS domain and uses EEG measurement in an interference paradigm as well as additional self-reports. While the current protocol focuses on the main study; relevant information on the EEG-substudy is also included.

A “mini RDoC” assessment battery developed in preliminary work [16]; [17] is used to assess the six RDoC domains: 1) Positive Valence System, 2) Negative Valence System, 3) Cognitive Systems, 4) Social Processes, 5) Arousal and Regulatory Systems, and 6) Sensorimotor Systems. Participants use a tablet computer to provide self-reports and perform two cognitive paradigms. They will also answer questions on demographic information (e.g., age, sex, gender, education, family background and social-economic status), care-related, and clinical variables (e.g., psychiatric history, family history of psychiatric disorders). Furthermore, external ratings by the treating clinicians are obtained (e.g., suicidality, somatic diseases, illness severity) and relevant medical information is extracted from patient records (e.g., number of therapy sessions, medication, length of hospital stay).

### 2.2 Study Population

Individuals eligible for inclusion in the study are aged between 18 and 80 years. Two groups of participants are included in this study: A psychiatric patient group and a healthy control group.

Inclusion criteria for the psychiatric patient group are the ability to read, write and comprehend the German language, have normal or corrected-to-normal vision, have the willingness and ability to provide informed consent and being an in- or outpatient of the participating psychiatric hospitals at the time of recruitment. Following the transdiagnostic approach, we will not select a particular group of psychiatric patients based on diagnosis. Exclusion criteria for the psychiatric patient group are a known current diagnosis of an acute severe somatic disease (e.g., pneumonia), severe cognitive disorder (e.g., amnestic syndrome), mental disability, and being admitted to the hospital involuntarily. Further, patients are excluded if they have been on the psychiatric ward for more than two weeks at the time of recruitment.

Inclusion criteria for the healthy control group are the ability to read, write and comprehend the German language, have normal or corrected-to-normal vision, and have the willingness and ability to provide informed consent. Participants in the healthy control group are excluded if they report having a known current diagnosis of an acute severe somatic disease (e.g., pneumonia), severe cognitive disorder (e.g., amnestic syndrome) or mental disability.

Based on recommendations for conducting factor analyses, consideration of feasibility and power size calculations, we intend to recruit a sample size of *n* = 300. For the factor analysis, both participants in the patient group and the healthy control group will be included. Medium effect size *ρ*^*2*^ = .10 with an optimal statistical power (1-*ß* > .95) could be detected with *n* = 300, whereas a good statistical power (1-*ß* > .80) would allow to detect effect sizes up to *ρ*^*2*^ = .07 and sufficient statistical power would allow to detect effect sizes up to *ρ*^*2*^ = .06.

Participation in the EEG substudy is optional for all participants enrolled in the main study. We aim to include approximately 33% or *n* = 100 of all participants, comprising both patients and healthy controls. Participants must provide additional informed consent specifically addressing the EEG procedures and must meet predefined eligibility criteria (i.e., no diagnosis or history of a central nervous system disorder, severe hyperkinetic disorder, or any condition affecting the scalp).

### 2.3. Procedures/ Assessments

#### 2.3.1.

Recruitment Recruitment for the present study is ongoing, started in November 2022 and includes patients on the inpatient units or day treatment centers of two psychiatric hospitals in Brandenburg, Germany, as well as healthy individuals. Patients are contacted by the study team within the first week after hospital admission during their stay. They are informed that their decision about participation will not affect their treatment in any way. After verbal and written information about the study, sufficient time for reflection, and verbal and written consent to participate in the study, inclusion and exclusion criteria are checked by the study team and treating physician. Assessments for the main study (standardized questionnaires and cognitive paradigms) take place within the first two weeks following admission and are ideally completed within a single session, but may be split across two days if needed. Healthy individuals are mainly recruited among students of the Brandenburg Medical School receiving course credits; however, additional participants may be included from the general community to ensure sample diversity. Healthy individuals interested in participation are informed and, after their consent is obtained, checked for inclusion and exclusion criteria.

#### 2.3.2. Measures

##### Global Assessment of RDoC Domains, covariates, and clinically relevant and care-related data

###### Self-report measures

Participants complete questionnaires on a tablet PC to report sociodemographic and clinical information. Sociodemographic variables include gender (both assigned at birth and self-identified gender), age, and socioeconomic status, operationalized through net household income, occupational status, years of education, and highest educational qualification. Participants also report on the number of children they have, their marital or partnership status, and their social living arrangements (e.g., whether they live with parents, children, or roommates). Clinical variables comprise self-reported psychological diagnoses, psychiatric family history, indicators of illness chronicity, including age at onset and duration of previous psychiatric treatment, self-rated severity (analogue to the CGI), and family history.

Next, a battery of items of standardized, self-report questionnaires will be administered via tablets to assess all RDoC domains except for the CoS domain. This test battery is referred to as “mini RDoC” and was developed by the research group Social and Preventive Medicine at the University of Potsdam. The *mini RDoC* was specifically designed to capture five RDoC domains through dimensional psychological assessment. The version used in the present study is based on previous foundational work, in which an initial transdiagnostic “mini-RDoC” assessment was created and validated using confirmatory factor analyses across a large psychiatric sample [16]. Further analyses examined associations between RDoC domains and disease severity in anxiety and depressive disorders, supporting the clinical relevance of the selected constructs [17]. Compared to the earlier version of the test battery [16], the revised assessment reflects a conceptual and structural refinement. Drawing on findings from the confirmatory factor analysis, we reorganized and expanded the allocation of constructs, particularly within the PVS, NVS, and SoP domains. Additionally, two new RDoC domains— Social Processes (SP) and Arousal and Regulatory Systems (ARS)—were systematically integrated into the updated version. These modifications led to a refined item-to-construct mapping (see Table 1), ensuring more systematic coverage of both valence-related and regulatory dimensions of mental functioning.

**Table 1.**
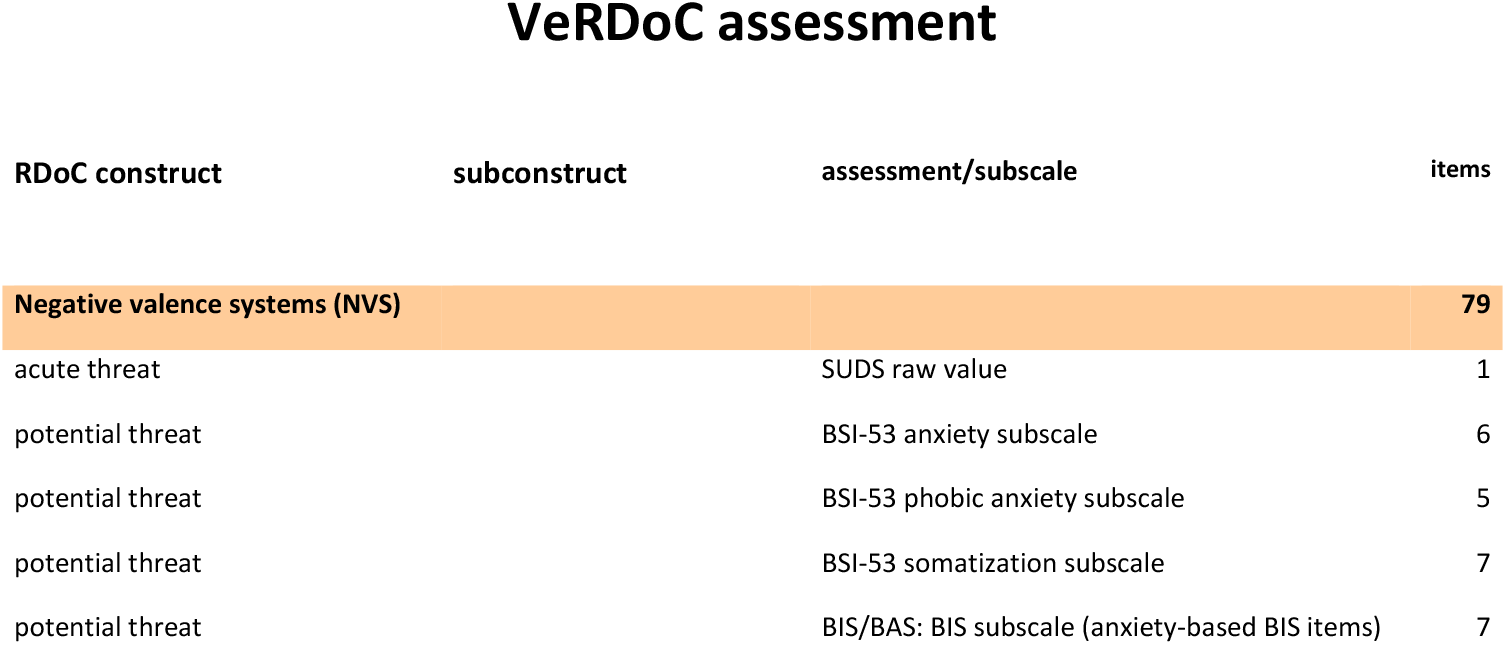
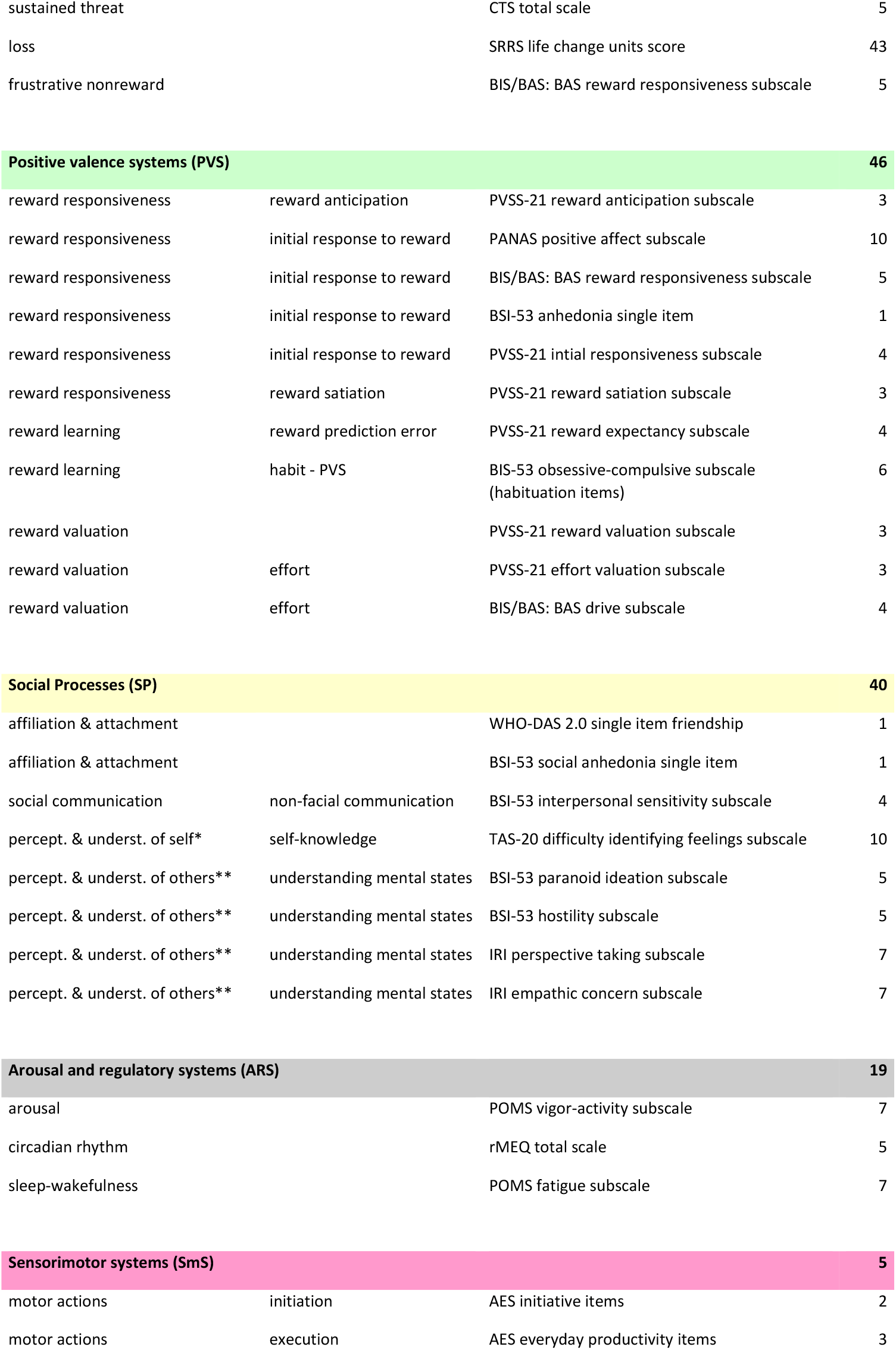
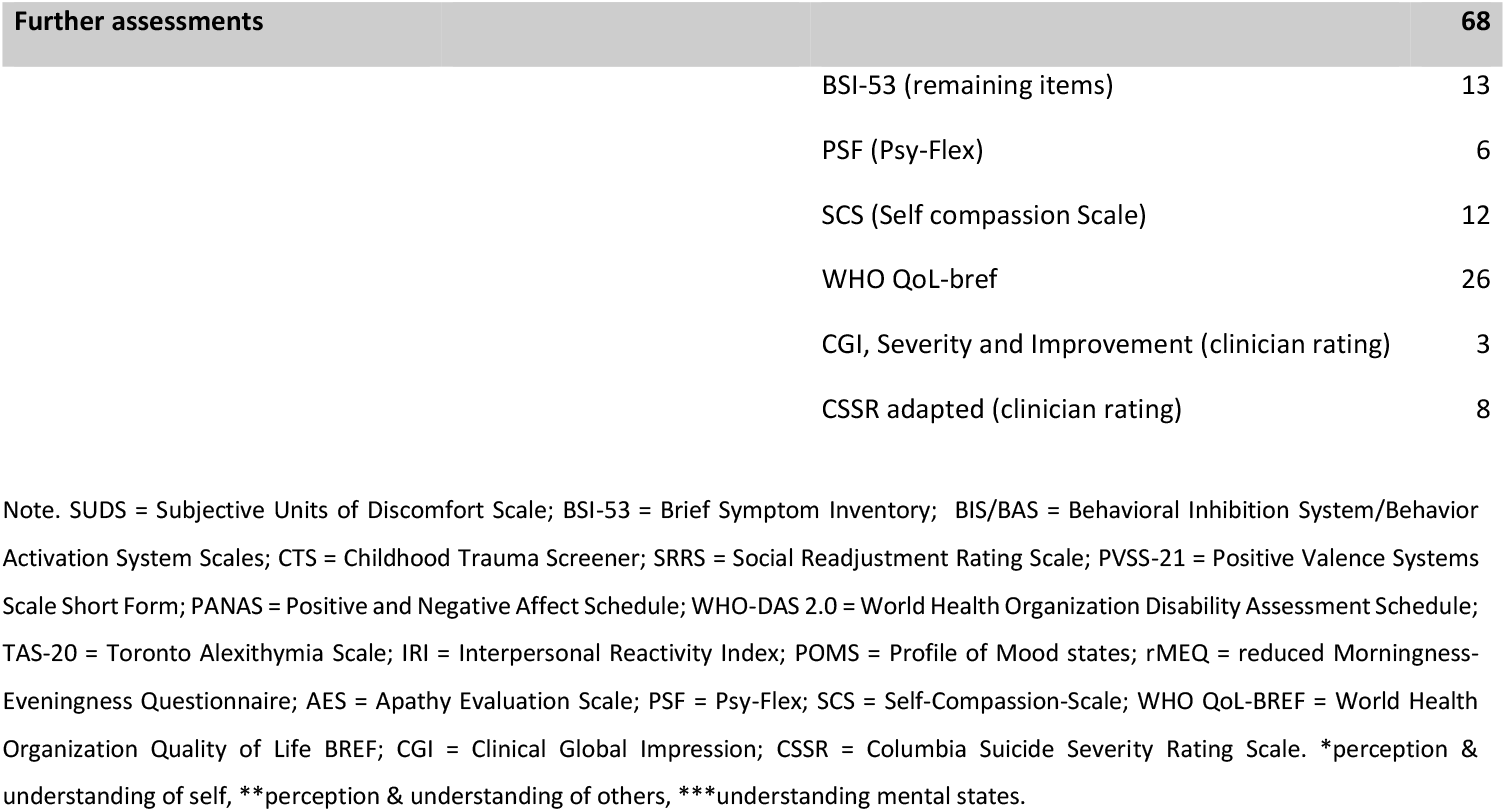
List of all VeRDoC Self-Report Assessments.

Finally, several standardized instruments are administered to assess broader aspects of mental health, psychological functioning, and well-being. The World Health Organization Quality of Life Questionnaire – Brief Version (WHOQOL-BREF; [18]) are administered to assess subjective quality of life across four domains, including physical health, psychological well-being, social relationships, and environment. It also includes one facet on overall quality of life and general health. The Psy-Flex Scale [19] is used to assess psychological flexibility [20], which is a core construct in Acceptance and Commitment Therapy (ACT). The short form of the Self-Compassion Scale (SCS; [21]) is used, which assesses trait levels of self-compassion and is comprised of the six subscales self-kindness, self-judgment, common humanity, isolation, mindfulness, and over-identification. Suicidality is assessed using a clinician rating of an adapted version of the Columbia Suicide Severity Rating Scale (C-SSRS; [22]). This tool provides a standardized measure of suicidal ideation and behavior and will be rated by patient’s treating clinicians. The Clinical Global Impression (CGI; [23]) scale will be used to evaluate severity of psychopathology and changes in severity over time. Ratings will include illness severity at admission (CGI-severity, CGI-S) and at discharge (also CGI-S) as well as overall improvement (CGI-Improvement, CGI-I), which indicates the change relative to baseline/admission. All ratings are completed by the treating clinician.

##### Care-related and clinical information from patient records

Other care-related and clinical information are extracted from electronic patient records. These include psychiatric and somatic diagnoses, information on pharmacological treatment, including current medication status and any recent changes made during the inpatient stay. Data on therapeutic interventions such as the number and type of group therapy/individual sessions attended are also extracted. The total duration of the current hospital stay is recorded, along with the time elapsed since the most recent inpatient stay, and whether there was another readmission within six months of the index stay. Finally, the reason for discharge is recorded, with a distinction made between regular and irregular discharges.

##### Cognitive Paradigms

Two tasks from the Cambridge Neuropsychological Test Automated Battery (CANTAB) are administered via tablet to assess the CoS domain: the Spatial Span Reverse 2.0 [24] and the Stop Signal Task [25]. The Spatial Span task assesses visuo-spatial working memory by presenting a sequence of flashing squares on the screen, which participants must remember and then reproduce in reverse order. The sequence length increases progressively until participants make two consecutive errors at the same level. The primary outcome is the longest sequence length correctly recalled in reverse order, reflecting working-memory capacity. The Stop Signal Task evaluates response inhibition by asking participants to respond to the direction of either a left- or right-headed arrow, while occasionally requiring them to withhold their response upon occurrence of an auditory stop signal. The CANTAB version employs an adaptive procedure in which the stop-signal delay is dynamically adjusted based on the participant’s performance. This approach maintains a roughly 50% inhibition success rate and allows for reliable estimation of the stop-signal reaction time, the primary outcome of inhibitory control.

##### Electroencephalographic Recordings and Cognitive Control Questionnaires within the EEG Substudy

An established interference paradigm, the *temporal flanker task* (e.g., [26–28]), that assesses several components of the event-related potential related to cognitive control (e.g., conflict-related N2, error-related negativity) as well as behavioral measures of conflict adaptation (e.g., reaction time, accuracy) is applied. Resting state EEG activity is also assessed, while participants are seated comfortably in a chair, let their minds wander, and alternately keep their eyes closed (5 minutes) and open (5 minutes) without falling asleep. Finally, the auditory mismatch negativity, which is an event-related potential elicited when the brain detects an irregularity in a stream of auditory stimuli, is assessed (e.g.,[29]). Here, we employ a simple oddball paradigm comprising frequency deviants (∼10% probability, 90 stimuli) and duration deviants (∼10% probability, 90 stimuli), embedded in a stream of standard tones (∼80% probability, 740 stimuli). Stimuli will be presented while participants watch a silent video.

In order to assess perceived daily cognitive control, participants completed the German versions of the Cognitive Failure Questionnaire (CFQ-G; [30]; [31]), the Urgency, Premeditation, Perseverance, Sensation Seeking Impulsive Behavior Scale (UPPS; [32], the Barratt Impulsiveness Scale version 11 (BIS-11; [33, 34]), the Obsessive Compulsive Inventory - Revised (OCI-R; [35]), and the Test of Attentional Style (TAS; [36]). As a covariate, the Edinburgh Handedness Inventory (EHI; [37]) is administered.

This part of the study takes place in a separate session and lasts approximately two hours, including EEG preparation, recordings, and the completion of questionnaires. The detailed design, methodology, and analysis plan are not within the scope of the current protocol and will be described in separate future publications. Importantly, the participation in the EEG session does not influence the procedures or outcomes defined in the main study.

### 2.4 Statistical Analyses

We intend to determine factor scores for the RDoC domains by conducting a confirmatory factor analysis. We will then analyze potential associations of RDoC variables and clinically relevant, care-related, and sociodemographic variables using multiple regression analyses.

#### 2.4.1 Confirmatory Factor Analysis (CFA)

To examine the latent RDoC factor structure of the mini RDoC assessment, a CFA will be conducted using maximum likelihood (ML) estimation. The analysis will be performed in R using the lavaan package [38] within RStudio [39]. Full information maximum likelihood (FIML) will be applied to account for missing data, under the assumption that data are missing at random (MAR). Latent variables will be standardized by fixing their variance to one, allowing all factor loadings to be freely estimated. To assess model fit, the proposed five-factor model will be compared to a unidimensional model in which latent variables are perfectly correlated (correlation set to 1), and to an orthogonal model assuming no covariance between latent variables.

Prior to model estimation, the distribution of indicator variables will be evaluated using the Shapiro– Wilk test. In case of significant deviations from normality, appropriate data transformations will be applied: right-skewed variables will undergo natural logarithmic (ln) transformation, while variables with large ranges will be normalized using the Johnson transformation. Variables that are already z-standardized, as well as dichotomous or categorical items, will not be transformed.

#### 2.4.2 Regression analyses

We then will conduct multiple linear regression analyses to examine the associations between the CFA derived RDoC factor scores and our measures of clinically relevant (e.g., psychological distress, quality of life, symptom severity) and care-related variables (e.g., length of hospital stay, readmission). For binary outcomes (e.g., readmission), logistic regression analyses will be used. To analyze whether sociodemographic and socioeconomic factors (e.g., age, gender, education, income) play a role, moderated regression models that include the interaction terms between the RDoC factor scores and the moderating variables (e.g., *PVS × Age*) will be employed. When modeling interaction effects involving categorical moderators (e.g., diagnostic group, gender), variables will be dummy-coded appropriately. Significant interactions will be further examined using simple slopes analysis and plotted to aid interpretation.

To analyze whether RDoC domains provide incremental predictive value beyond traditional diagnostic categories and illness severity, hierarchical regression analyses will be employed using a stepwise approach. Diagnosis and illness severity will be entered in the first step, followed by RDoC factor scores in the second step. The change in explained variance will be used to evaluate the additional contribution of RDoC constructs. For binary outcomes such as suicide attempt (yes/no) or readmission, hierarchical logistic regression will be used with a stepwise approach. Lastly, to investigate whether RDoC domains can predict clinical change (CGI-I scores), ordinal logistic regression will be employed. Statistical analyses will be conducted using SPSS [40] and a significance level of α = .05 will be used for all tests.

In addition to significance testing, effect sizes will be reported to allow interpretation of the magnitude and clinical relevance of the findings. For linear regression analyses, standardized regression coefficients (β) and their confidence intervals will be reported. For logistic and ordinal logistic regression models, odds ratios (*OR*) with 95% confidence intervals will be presented. Where applicable, change in explained variance (*ΔR*^*2*^) or pseudo *R*^*2*^ measures (e.g., Nagelkerke’s *R*^*2*^) will be used to quantify model improvement.

Although the current protocol focuses on the main study, certain additional measures collected in the embedded EEG substudy may be used for exploratory analyses to address specific secondary or additional research questions within the broader scope of the main study. These analyses can and will be conducted only in the substudy subsample and will be clearly identified as such. The limited sample size and potential selection bias will be taken into account in interpreting these results. Importantly, none of the primary or secondary endpoints of the main study rely on data collected exclusively in the substudy.

### 2.5 Patient and Public Involvement

This study did not involve patients or the public in the design, conduct, reporting, or dissemination plans.

## 3. Ethics and dissemination

The study was approved by the local ethics committee of the Brandenburg Medical School – Theodor Fontane (No. E-01-20220822) and is conducted in accordance with the Declaration of Helsinki. All participants receive written information about the study’s aims, procedures, and their rights, and are also verbally informed by trained study staff prior to providing written informed consent. As part of the consent process, participants agree that, in the event of acute psychological distress or suicidal ideation, study staff may inform the treating clinicians. Participants are informed that their participation is voluntary and that both participation and refusal to participate will not affect their treatment. All participants are given sufficient time to consider their participation, and both verbal and written informed consent is obtained prior to inclusion in the study. Participants are also informed of their right to withdraw at any time without consequences. This observational study involves minimal risk. Nonetheless, during all assessments study staff is present in the event that a participant reports acute psychological distress. If a participant shows signs of acute severe distress or reports suicidal thoughts, a predefined safety protocol is activated, which includes communication with the clinical care team on site and will occur promptly to ensure continuity of care and participant safety.

Data is stored securely in compliance with local data protection regulations and personal data is stored separately from assessment responses. All data are pseudonymized to ensure participant confidentiality.

Study results will be disseminated through publication in peer-reviewed journals and presentations at scientific conferences. Study materials (e.g., translated assessment instruments, analysis code for published results) will be deposited in an open-access repository and updated continuously (https://osf.io/u9fpw; [41]) to promote transparency and reproducibility.

## Author Contributions

CRediT: KKS: Conceptualization, Data curation, Funding acquisition, Investigation, Methodology, Project administration, Supervision, Writing – original draft, Writing – review & editing; JM: Conceptualization, Data curation, Funding acquisition, Investigation, Methodology, Project administration, Writing – review & editing; KJ: Conceptualization, Funding acquisition, Methodology, Supervision, Writing – review & editing; FW: Project administration, Investigation, Visualization, Writing – review & editing; JR: Conceptualization, Data curation, Methodology, Writing – review & editing; BRF: Conceptualization, Data curation, Methodology, Resources, Writing – review & editing; MH: Resources, Writing – review & editing; JB: Conceptualization, Funding acquisition, Resources, Methodology, Supervision, Writing – review & editing; MAR: Conceptualization, Funding acquisition, Resources, Supervision, Writing – review & editing; MT: Conceptualization, Data curation, Funding acquisition, Methodology, Project administration, Resources, Supervision, Writing – original draft, Writing – review & editing. All authors have read and approved the manuscript. Each author agrees to be accountable for all aspects of the work, ensuring that questions related to the accuracy or integrity of any part of the work are appropriately investigated and resolved.

## Funding statement

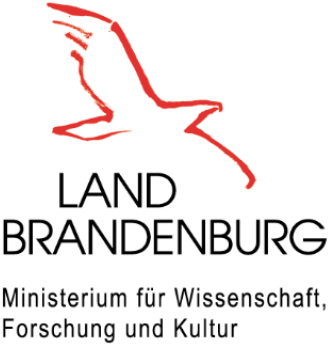

This project was funded by the Ministry of Science, Research and Cultural Affairs of the State of Brandenburg.

## Competing interests statement

The authors declare no competing interests.

## Acknowledgements

None.

## Data Availability Statement

The datasets generated and/or analyzed during the current study are not publicly available due data security restrictions but are available from the corresponding author on reasonable request.

## Notes

### Competing Interest Statement

The authors have declared no competing interest.

### Author Declarations

Ethics committee of the Brandenburg Medical School - Theodor Fontane gave ethical approval for this work.

